# Ten Years of Clinical Negligence Claims in Otolaryngology: Trends, Costs, and Preventable Harm in the UK National Health Service

**DOI:** 10.1101/2025.05.03.25326930

**Authors:** Behrad Barmayehvar, Christopher Metcalfe, Jameel Muzaffar

## Abstract

**Introduction:** With greater pressures on the British National Health Service (NHS), litigation is increasing across surgical specialties. The national litigation trends in otolaryngology have not been adequately explored in the recent years, particularly in the aftermath of the COVID-19 pandemic.

**Objectives:** The primary objective was to analyse NHS litigation data in otolaryngology over the last decade, exploring the impact of the pandemic on claim volumes and costs. The secondary objective was to identify common areas of concern in patient care.

**Methods:** National-level anonymised negligence claims data for the financial years 2013/14 to 2023/24 were obtained from NHS Resolution via a Freedom of Information request. The data included claim status, claimant’s age group, incident and notification dates, primary cause and injury codes, and the associated costs.

**Results:** A total of 1,450 claims were received and 1,594 cases were closed. Number of received claims declined from 2020/21 onwards, whereas, total costs followed the closed claim numbers, both gradually increasing with spikes in 2018/19 and 2022/23. The total costs were also driven up by a sharp increase in damages and steadier increase in claimant legal costs. Of the closed cases, 58% resulted in damages payment. The most frequent claim cause was delay in treatment or diagnosis, whilst inadequate intra-operative monitoring led to the highest mean payout. The most frequent injury code was unnecessary pain, whilst brain damage, although rare, was the costliest. We identified six Never Event categories, with foreign body left in situ being the most common.

**Conclusion:** Whilst received claims are declining since the pandemic, litigation costs in otolaryngology continue to rise. The results highlight areas of care for targeted improvement, including earlier diagnosis and treatment, robust monitoring, and better communication to reduce preventable harm.

## Introduction

Over the past two decades, the British National Health Service (NHS) has experienced a substantial increase in litigation costs. According to the National Audit Office, from 2006/7 to 2016/17, annual spending on clinical negligence quadrupled from £0.4 billion to £1.6 billion [1]. This upward trend has continued, with damages for settled clinical negligence cases reaching £2.8 billion in 2023/24, an increase of 75% since 2016/17 [2].

NHS Resolution (NHSR) is a public body with many functions, including managing clinical and non-clinical claims on behalf of the NHS and promoting patient safety [3]. The Clinical Negligence Scheme for Trusts (CNST) is the current NHSR framework to fund litigation costs. Although it is voluntary, all NHS Trusts currently participate in this scheme, and their collective annual membership funds provide the financial support for any litigation cost within NHS [4]. In 2024, NHSR reported “welcome positive signs” that the rate of increase of damages was dropping [2]. This is largely due to NHS Resolution’s efforts to collaboratively resolve disputes without needing formal court proceedings, which was achieved in a record 81% of cases in 2023/24, an increase of 4% since 2021/22 [2,5]. However, the financial burden remains high as overall damages and claimant legal costs continue to soar [2], highlighting the need for effective strategies to improve patient safety and satisfaction.

Whilst the majority of claims are related to surgical specialties, maternity care consistently has the highest litigation costs, averaging £2.6 million per claim [6]. Otolaryngology, as a surgical specialty, has experienced a notable increase in litigation over the past two decades [7,8]. Between 1996 and 2017, there were 1,952 claims against otolaryngology departments, resulting in payouts exceeding £108 million [8]. A focused analysis of 727 otolaryngology-related claims between 2013 and 2018 in conjunction with Getting It Right First Time data revealed that otolaryngology had the 7th highest claim volume out of 16 surgical specialties [9]. Head and neck surgery accounted for the highest number and cost of claims, followed by otology and rhinology. Notably, over half of these claims were related to surgical procedures, emphasising the critical need for meticulous operative practices and informed consent processes [9].

The COVID-19 pandemic further complicated the landscape of otolaryngology practice. The pandemic led to the halt or delay in the activity of many elective clinic and operative sessions and led to increased waiting times [10,11]. Moreover, the considerable risk of aerosol generation during otolaryngology procedures necessitated rapid adaptations in surgical protocols [12,13]. Until now, post-pandemic NHS litigation trends in otolaryngology have not been rigorously examined.

This study aims to analyse clinical negligence claims reported to NHSR between the 2013/14 and 2023/24 fiscal years with a view to assess the impact of the recent increase in dispute resolutions without legal proceedings and the COVID-19 pandemic. The primary objective was examining the trends in claim volumes, primary causes, associated injuries, and financial outcomes; whilst the secondary objective was identifying areas for potential intervention to enhance patient care.

## Materials and Methods

A Freedom of Information (FOI) request was submitted to the NHSR to obtain relevant data on the clinical claims handled by this body between 2013/14 to 2023/24. The data were derived from three different current and historic schemes that have managed the negligence claims in the NHS: Clinical Negligence Scheme for Trusts (CNST), Existing Liabilities Scheme, and Department of Health and Social Care (DHSC) Clinical Liabilities Scheme [14]. NHSR provided anonymised national-level data on the volume of received and closed claims, age group of claimants, date of incident taking place and the claim being notified, primary cause and injury codes, and lastly costs of closed claims. Children were defined as aged 0 to 17, whilst adults were 18 years or more by NHSR. Data was analysed and visualised with Microsoft Excel (Microsoft Corp, Washington, USA) and R 4.5.0 (R Foundation for Statistical Computing, Vienna, Austria).

The total costs of closed claims were further broken down into damages, claimant legal costs and NHS legal costs. Periodical Payment Orders (PPOs) involve an arrangement where the claimant receives an initial lump sum followed by regular payments to cover their ongoing caring needs, often for life. The disclosed data included lump sum compensation, legal expenses, and PPO payments made to the end of the settlement year, but future PPO payments that NHSR has committed to beyond that period were not included. In cases without PPOs, financial data was reported up to the end of the closure year for the case.

It is important to note that the received claims by NHSR during our 10-year period of interest differ from the closed claims for the same period; the former are newly notified incident claims, whilst the latter are cases that are settled either with or without damages. This is because some notified claims may only be closed at a much later date with significant delays between an incident being reported and the claim being resolved. Open claims are not included in the NHSR reports usually as they are subject to frequent change. Information on categories with less than five claims were withheld under the Section 40(3A) of the FOI Act [15], as there is a risk that individuals could be identified, either from the data alone or when combined with other publicly accessible information.

Never Events are defined as “serious incidents that are wholly preventable because guidance or safety recommendations that provide strong systemic protective barriers are available at a national level” [16]. NHS Improvement published an updated list of Never Events in February 2021 [17]. We used this list to identify claims that could potentially relate to a Never Event based on their primary cause or injury. Further descriptive details on the circumstances leading to each case are not allowed to be released via FOI requests as they may identify individuals involved in the claims.

## Results

### Received claims

Data was requested on 15th December 2024 and received in batches on 28^th^ January 2025 and 13^th^ March 2025. The total number of received claims were 1,450 between 2013/14 and 2023/24. The number of received claims in fact had a gradual decline from the year 2020/21 onward, dropping from 130 received claims in 2020/21 to 106 in 2023/24 (Figure 1). Most of the received claims pertained adult claimants (1,267, 87%), whilst the remaining cases were either related to children (150, 10%) or had unknown age at the time of notification (33, 2%).

**Fig 1.**
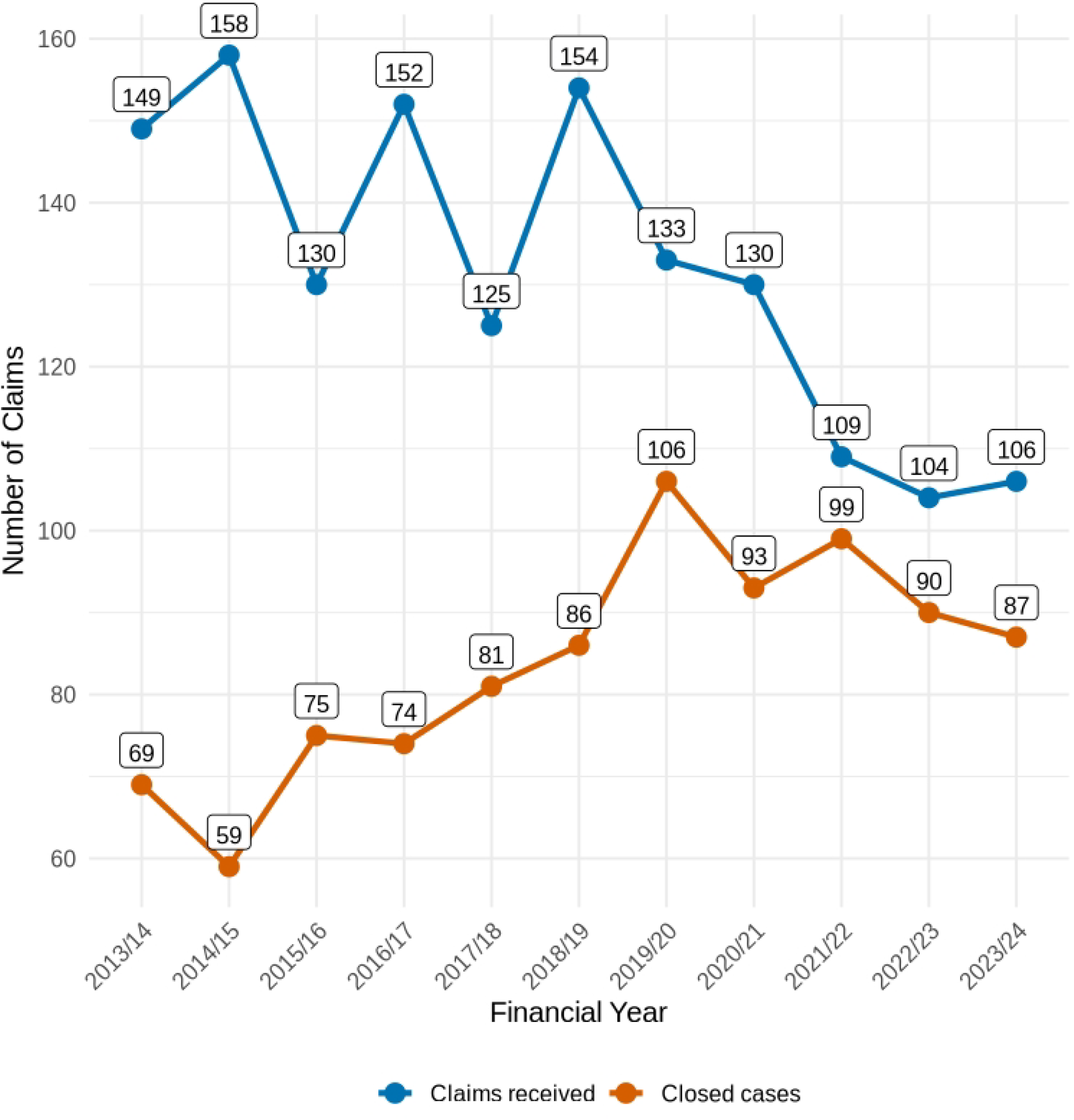
Number of Claims Received and Closed per Financial Year (2013/14–2023/24) This line chart shows the trends in the number of clinical negligence claims received and closed per year by NHS England. Data are stratified into received (blue line) and closed (orange line) claims. Overall, the number of claims received has declined since 2020/21, whereas the number of closed claims peaked in 2019/20.

### Closed claims

In total, 1,594 claims were closed during this period. The number of yearly closed claims show a general rise with a peak in 2019/20 (Figure 1). Most of these claims (919, 58%) were settled with damages payments, but 675 (42%) claims were closed without damages, notwithstanding the NHS legal costs. Of the 919 claims settled with damages, 811 (88%) belonged to adult claimants, whilst 98 (11%) were paediatric cases and 10 (1%) remained of an unknown age at the notification time.

The total costs generally reflected the trend of the yearly volume of closed claims. Breaking down the costs further revealed that the damages payment and volume of closed claims were the driving forces behind the sharp rises seen in the total costs. Legal costs to both the NHS and claimant also had an upward trend, but their rate of increase, and hence contribution to the rise in total costs, were more modest than that seen in damages. There were two significant peaks in damages and total costs in 2018/19 and 2022/23, which were disproportionate to their respective closed claims volume.

Delving into the incident years when the closed claims were originally received revealed that a few claims (less than 5, hence not specified) dated as far back as 1981/82, whilst the most recent incident year was 2021/22. The three incident years that had the highest volume of closed claims were 2011/12, 2012/13, and 2015/16 with 94, 92 and 93 closed claims, respectively. In contrast, 2012/13 and 2015/16 incident years had the greatest number of unsuccessful claims (79 and 71 respectively) that resulted in no damages payment. The total cost of closed cases for those received in the 2011/12 incident year was the highest at about £21 million; however, 2012/2013 incident year with a similarly high volume of closed cases only recorded £11 million total cost. This demonstrates that the volume of cases does not necessarily reflect the complexity and total costs of the cases.

**Fig 2.**
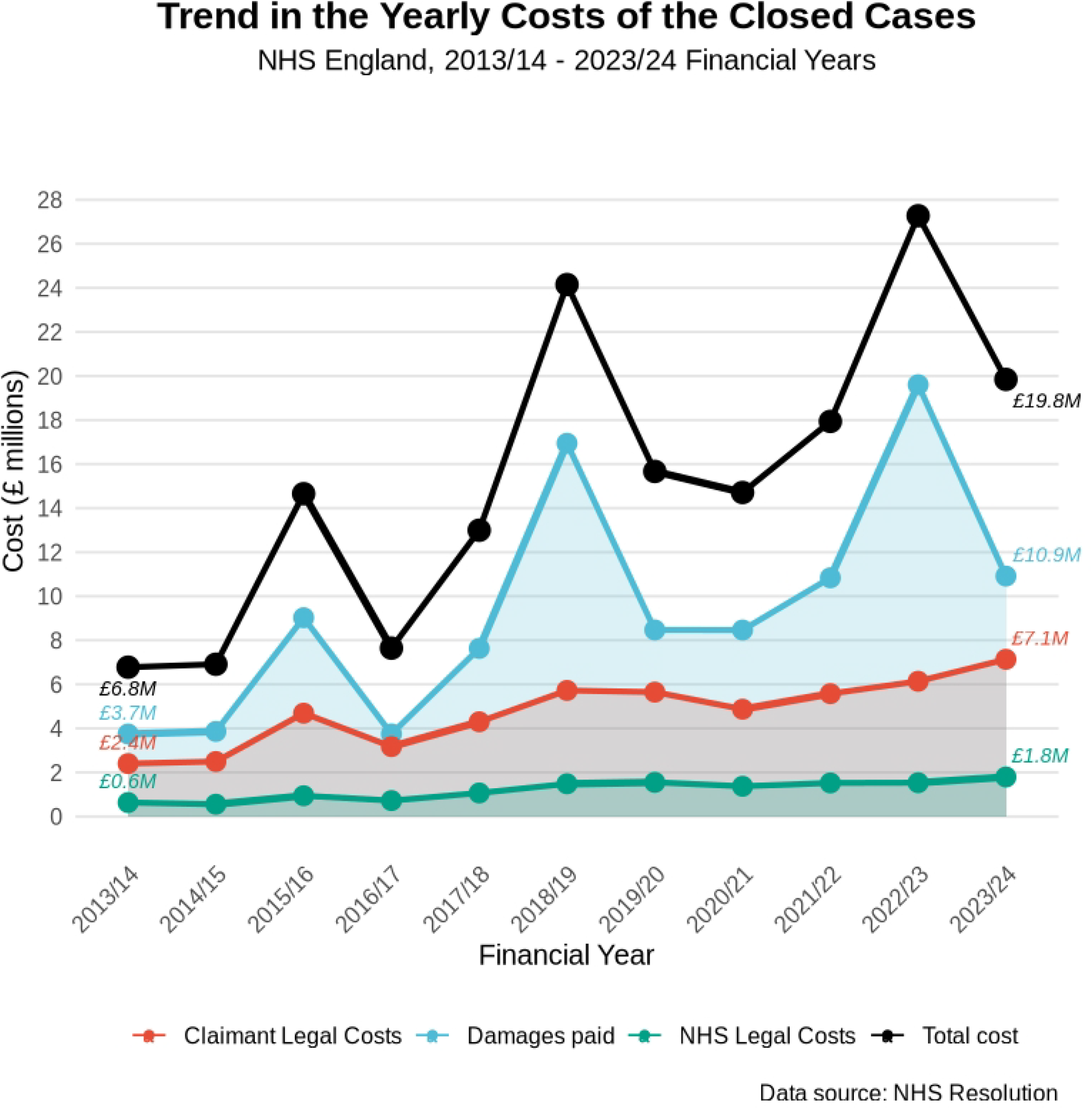
Trend in Yearly Costs of Closed Claims (2013/14–2023/24) Stacked area and line chart illustrating the breakdown of costs associated with closed clinical negligence cases in otolaryngology. Total costs (black line) are decomposed into claimant legal costs (red), NHS legal costs (green), and damages paid (blue). Notable peaks occur in 2018/19 and 2022/23 due to high damages payouts.

### Primary cause analysis

Failure or delay in treatment and diagnosis dominate most of the closed claims with 188 and 186 claims, respectively. These two causes alone account for over £78 million in total costs. Failure to warn is noteworthy for its relatively high payouts whilst remaining a largely preventable cause that relies on effective communication and consent. Inadequate intra-operative monitoring stands out as it has the highest mean payout per claim, suggesting profound consequences such as long-term disability may have ensued from these incidents. In order to allow a direct comparison with the work of Patel et al, we have provided a list of the twenty most frequent primary causes and injuries of the period along with their associated costs in Table 1.

**Table 1.** The top 20 most frequent primary causes and injuries along with their associated total costs as reported for the closed claims between 2013/14 and 2023/24.

| Primary Cause | No. of Claims | Total Paid (£) | Primary Injury | No. of Claims | Total Paid (£) |
| --- | --- | --- | --- | --- | --- |
| Fail / Delay Treatment | 188 | 36,611,071 | Unnecessary Pain | 163 | 10,176,931 |
| Failure/Delay Diagnosis | 186 | 42,053,022 | Additional/ Unnecessary Operation(s) | 125 | 8,538,827 |
| Intra-Op Problems | 84 | 10,330,408 | Nerve Damage | 75 | 12,651,877 |
| Fail To Warn or Informed Consent | 82 | 18,993,124 | Cancer | 73 | 13,029,809 |
| Inappropriate Treatment | 65 | 9,007,768 | Deafness | 62 | 9,589,375 |
| Operator Error | 51 | 6,206,012 | Advanced Stage Cancer | 51 | 15,311,099 |
| Failure To Perform Tests | 29 | 3,385,307 | Fatality | 43 | 10,115,627 |
| Fail To Recognise<br>Complication Of | 27 | 5,152,269 | Partial Hearing Loss | 33 | 6,024,034 |
| Foreign Body Left In<br>Situ | 23 | 504,802 | Cosmetic<br>Disfigurement | 26 | 2,083,360 |
| Fail To Follow-Up<br>Arrangements | 17 | 1,708,956 | Brain Damage | 26 | 42,839,552 |
| Inadequate Nursing<br>Care | 15 | 514,668 | Scarring | 24 | 1,209,698 |
| Delay In Performing<br>Operation | 11 | 572,749 | Perforation | 20 | 1,439,188 |
| Fail To Act On<br>Abnormal Test Results | 11 | 2,471,905 | Psychiatric/<br>Psychological Damage | 18 | 2,404,448 |
| Wrong Site Surgery | 11 | 348,569 | Vocal Cord Damage | 17 | 3,903,105 |
| Medication Errors | 9 | 1,279,534 | Other Infection | 15 | 795,374 |
| Failure To Perform<br>Operation | 9 | 3,772,311 | Other Visual Problems | 15 | 2,313,181 |
| Diathermy<br>Burns/Reaction To<br>Prep | 8 | 331,738 | Burn(s) | 15 | 289,990 |
| Performance Of<br>Operation Not<br>Indicated | 8 | 837,335 | Other | 11 | 1,474,703 |
| Inadequate Monitoring<br>Intra-Op | 8 | 8,242,207 | Blindness | 9 | 1,548,980 |
| Lack Of<br>Assistance/Care | 7 | 849,200 | Dental Damage | 7 | 102,842 |

### Primary injury analysis

The highest number of injuries recorded were due to unnecessary pain (163 claims) and unnecessary operations (125 claims). Brain damage stands out due to its highest mean cost per injury despite having only 26 claims. Cancer-related claims (combined early and advanced stages) also represent a considerable portion of high-value payouts, reflecting the devastating impact of delays in diagnosis and treatment of cancers on the patients. Auditory injuries (deafness and partial hearing loss) together account for £15.6M across 95 claims, which are likely to be uniquely high in otolaryngology compared to other specialties. These claims could arise from the inherent risks in otology surgery, ototoxic drug administration or delayed treatment of reversible hearing losses. Surprisingly, cosmetic disfigurement and scarring injuries are relatively low in frequency and cost, possibly due to better mitigation or patient expectation management (Table 1).

### Never Events

Six Never Event categories were identified; five based on the primary cause coding and a further Never Event from injury codes. Table 2 shows the frequency and costs of each Never Event. The most frequent Never Event was foreign body left in situ (23 claims), whilst the costliest was error with agent route selection category. Wrong Site Surgery had a surprisingly low payout for the gravity of its preventable error (£348,569), possibly due to quick recognition and mitigation of harms.

**Table 2.** Frequency and cost of Never Event categories in otolaryngology identified through the primary cause and injury reports of NHSR between 2013/14 to 2023/24. The categories with less than 5 recorded claims are censored by NHSR due to data protection laws.

| Never Event | Number<br>of<br>Claims | Damages<br>Paid (£) | NHS Legal<br>Costs (£) | Claimant<br>Legal<br>Costs (£) | Total Paid<br>(£) | Mean cost<br>per claim<br>(£) |
| --- | --- | --- | --- | --- | --- | --- |
| Foreign Body<br>Left In Situ | 23 | 167,688 | 30,914 | 306,200 | 504,802 | 21,948 |
| Burn injuries | 15 | 113,673 | 19,605 | 156,713 | 289,990 | 19,333 |
| Wrong Site<br>Surgery | 11 | 172,821 | 13,061 | 162,687 | 348,569 | 31,688 |
| Error With<br>Agent/<br>Dose/<br>Route/<br>Selection | 6 | 165,300 | 24,032 | 116,240 | 305,572 | 50,929 |
| Misplaced<br>Naso/<br>orogastric<br>tube | <5 | censored | censored | censored | censored | N/A |
| Retained<br>Instrument<br>Post-Operation | <5 | censored | censored | censored | censored | N/A |

## Discussion

The key finding of our work is that despite a decline in the number of received claims since 2020/21, total financial payouts have continued to soar. This rising cost appears to have been driven by significant increases in damages and, to a lesser extent, by the gradual rise in claimant legal costs and volume of closed cases. Claimant legal fees consistently exceeded NHS legal fees, often by a large margin. This pattern could suggest that claims are becoming increasingly complex with associated heavier costs.

The COVID-19 pandemic led to “missing patients” not being referred to specialties [11] or being lost to follow up [18]. This could have been due to various factors, such as seeking private healthcare or avoiding healthcare settings altogether due to fear of infection risk [19]. We speculate that these “missing patients” could have had delayed diagnoses or presented with complex advanced diseases that were more difficult to manage post-pandemic and hence led to more complex and costly litigation. This could also partly explain the high number of claims with delayed diagnosis and treatment as their primary cause.

On the other hand, the reduction of received claims post-pandemic could be attributable to several factors. A more sympathetic attitude by the public towards NHS workers and their sacrifices during the pandemic [20] could have led to less patients deciding to take legal action against NHS staff. Furthermore, the new Strategy of NHSR for 2022 to 2025 emphasised greater focus on settling disputes without escalation to formal legal proceedings, which has prevented costly legal proceedings in up to 81% of disputes in 2023/24. On the other hand, public satisfaction with NHS services has plummeted since 2020, and is at its lowest since the since the first British Social Attitudes (BSA) survey in 1983 [21]. This is of great concern and might translate into future escalation in both numbers and costs of litigations.

Delving into the causes and injuries underlying claims offers insights in a number of areas of patient safety and care quality. Failure or delay in treatment and diagnosis were the most frequent claim causes, costing over £78 million in damages. Both can arise for a variety of reasons; some can be directly influenced by complexity of cases and delay in recognition of signs, but others may arise from systemic factors outside of clinicians’ control such as the pandemic [22] or the ongoing staffing and resource shortages [23]. Inadequate intra-operative monitoring was the costliest primary cause, and arguably, the upfront cost of setting up equipment and systems for improved patient safety can mitigate the future costs of such claims. For example, intra-operative monitoring of facial or recurrent laryngeal nerves, endoscopic sinus surgery navigation systems and perhaps, validated Artificial Intelligence tools of the future [24], can all play a role in minimising the occurrence of claims related to poor monitoring alone.

The identification of Never Events, although relatively rare, highlights preventable safety lapses such as foreign bodies being left in situ. While the associated payouts were relatively low, their occurrence raises concerns about surgical safety protocols. Adherence to the evidence-based World Health Organization (WHO) checklist is an important initial step in reducing errors related to monitoring and preventable Never Events [25].

With respect to reported injuries, unnecessary pain and procedures were the most common. These may suggest system-level procedural or communication failings that lead to unwanted patient suffering and avoidable invasive treatments, respectively. Furthermore, although brain damage was relatively rare (26 cases) amongst closed claims, it was disproportionately costly, contributing to nearly 20% of the total financial burden.

Our results generally align with other published literature, except our finding of decline in number of received claims since 2020/21, which we have not found reported in similar studies. Both volume and cost of claims have shown an upward trend in most specialties in the last two decades [6,26]. Between 2004 and 2014, failure or delay in treatment and diagnosis and failure to warn/adequately consent were the three commonest causes for claims across 11 surgical specialties [26]. The same top causes were identified in otolaryngology with the addition of intraoperative problems in the decade preceding our study (2003 to 2013) [7].Our findings on injuries are comparable to other specialties; brain and neurological injuries rank amongst the top three costliest of injuries in general surgery, orthopaedics and obstetrics [27–29]. This is likely due to the grave loss of quality of life and lifelong dependency that often results from such injuries.

### Limitations

There are some limitations to conclusions that can be drawn from NHSR reports. Firstly, the data show some fluctuation in the number of received and closed claims each year, along with their associated costs. These variations reflect the diverse nature of individual cases managed by NHSR which can vary significantly. Complex and costly cases can skew the cost figures for some years substantially, even if the overall number of claims decrease. Therefore, such fluctuations should be interpreted with caution.

Secondly, claims reported or received in a given year often correspond to incidents that took place many years earlier. Due to the complexity of clinical negligence cases and the extensive investigations required to reach a resolution, submitted claims may take several years to be settled and officially closed. This time lag can give seemingly different cost figures even if the volume of received claims drops.

Thirdly, because of the way data is extracted, a single claim may appear multiple times in the dataset across different years. This could happen if a case initially closed without any damage payment is subsequently challenged, reopened, and then closed again upon final resolution.

We did not have the data to compare other specialties or individual Trusts in the same period. Similarly, we were not able to break down the claims by the subspecialties within otolaryngology, nor could we ascertain which claims were related to claimants who had undergone a surgical procedure. Patients and public were not directly involved in the design and analysis of the study, and we would recommend them to be involved in future qualitative studies.

### Ethics Statement

This study used anonymised, publicly available data provided by NHS Resolution following a Freedom of Information request. As the dataset did not contain any identifiable personal information, ethical approval was not required.

## Conclusions

The analysis of otolaryngology litigation claims over the last decade shows that despite a reduction in new claims in recent post-pandemic years, the increasing financial burden on the NHS remains a significant concern. Delay in diagnosis and treatment along with intra-operative monitoring failures, constitute the most frequent and costly primary causes respectively, pointing to key areas for intervention. Developing streamlined diagnostic and treatment pathways, investing in intra-operative safety technologies and surveying patients’ views on our services are all potential strategies to reduce harm and litigation risk.

As unnecessary pain and procedures were the most frequent reported injuries, otolaryngologists should ensure their consenting process is well-informed and meticulous, whilst all healthcare professionals involved in the patients’ journey, whether in outpatients or inpatients, should maintain high standards of communication. Policy makers and local health authorities can minimise barriers to effective communication by providing adequate staffing, training, and consultation time. Hospitals should maintain a structured approach to incident analysis, with a fresh focus on Never Event prevention strategies.

Lessons from this study should be integrated into ongoing patient safety initiatives, training programs, and policy reforms to ensure that patient outcomes improve while mitigating the growing legal and financial pressures facing the NHS. Future qualitative studies should focus on views of a sample claimants and healthcare professionals to gain a better understanding of barriers in communication and optimal outcomes.

## Acknowledgments

We would like to sincerely thank the NHS Resolution team who provided the data in a clear format and constructively clarified our queries about the data.

## Supporting information captions

## Author Contributions

BB: Conceptualisation, formal analysis, investigation, project administration, visualisation, writing of original draft. CM and JM: Methodology, resources, supervision, and validation. BB, CM and JM: Review and editing.

## Data Availability Statement

The data are publicly available upon reasonable request from the NHS Resolution public body.

